# Influenza’s Economic Burden and the Impact of Adult Vaccination

**DOI:** 10.64898/2025.12.23.25342685

**Authors:** Robert Popovian, Wayne Winegarden

**Affiliations:** Chief Science Policy Officer, Global Healthy Living Foundation, Senior Health Policy Fellow, Progressive Policy Institute, and Senior Visiting Health Policy Fellow, Pioneer Institute; Senior Fellow and Director of the Center for Medical Economics and Innovation at the Pacific Research Institute

## Abstract

**Background:** Seasonal influenza imposes a significant clinical and economic burden in the United States despite the availability of effective vaccines.

**Objectives:** To estimate the cost of illness associated with seasonal influenza among U.S. adults and to examine the relationship between vaccination coverage and influenza related outcomes.

**Methods:** We combined Centers for Disease Control and Prevention influenza burden estimates with contemporary healthcare utilization, cost, and labor market data to estimate direct medical costs and productivity losses for the 2023 24 influenza season. Panel data regressions with fixed effects were used to evaluate the association between adult influenza vaccination rates and hospitalization and mortality outcomes using data from the 2010 11 through 2023 24 seasons. Scenario analyses assessed how alternative vaccination rates would have affected costs and mortality.

**Results:** Influenza among adults was associated with an estimated $29 billion in total economic burden in the 2023 24 season, including approximately $16 billion in direct healthcare costs and $13 billion in productivity losses. Higher vaccination rates were significantly associated with lower mortality among adults aged 18 years and older and reduced hospitalization rates among adults aged 50 years and older. Achieving historical peak vaccination coverage would have reduced total costs by approximately $3 billion and averted more than 8000 deaths.

**Conclusions:** Adult influenza vaccination is associated with substantial reductions in mortality and economic burden, underscoring its value as a cost relevant public health intervention.

## Introduction

Influenza remains a substantial public health challenge in the United States despite the availability of annual vaccination.^1^ The Centers for Disease Control and Prevention (CDC) estimates that, in recent non-pandemic years, influenza has resulted in approximately 9.3–40 million symptomatic illnesses annually, 120,000–700,000 hospitalizations, and 6,300–51,000 deaths each year.^2^ The burden of disease is particularly pronounced during high-severity seasons.

The most recent finalized CDC data (2023-24) indicate that the influenza season was associated with an estimated 40 million symptomatic illnesses, 18 million medical visits, 470,000 hospitalizations, and 28,000 deaths.^3^ Beyond its clinical toll, influenza contributes substantially to the economic and societal burden of disease, resulting in billions of dollars in annual losses from direct medical costs and reduced productivity.

These figures underscore the broad reach of influenza, which imposes a heavy toll not only on morbidity and mortality but also on outpatient visits, hospital stays, lost productivity, and the strain on the healthcare system. Previous analyses have shown that influenza accounts for one of the highest annual cost burdens among vaccine-preventable illnesses in the U.S., making prevention a priority for health policymakers and healthcare systems alike.^4^

### Purpose of the Study

The purpose of this study is threefold:

1. Estimate the cost of illness associated with influenza utilizing the most recent data concerning incidence of disease, utilization of healthcare services, and the cost related to such services.
2. Examine the relationship between flu vaccination rates and burden of illness.
3. Approximate the economic impact of flu vaccination on the cost of illness associated with influenza.

By linking vaccination uptake to cost and outcome metrics, this study aims to generate actionable evidence for public health and policy decision-makers.

## Methods

### Study Design

This study employs a retrospective cost-of-illness framework combined with panel-data regression analyses to evaluate the economic burden of seasonal influenza among adults in the United States and to assess the association between influenza vaccination coverage and influenza-related outcomes. The analysis integrates publicly available epidemiologic, cost, and labor market data spanning multiple influenza seasons.

### Data Sources

Influenza burden estimates, including symptomatic illnesses, medical visits, hospitalizations, deaths, and vaccination coverage rates, were obtained from the Centers for Disease Control and Prevention (CDC). Seasonal estimates by adult age category (18–49 years, 50–64 years, and _≥_65 years) were drawn from CDC influenza burden and vaccination surveillance data for the 2010–11 through 2023–24 influenza seasons.

Healthcare cost inputs were derived from published peer-reviewed literature. All cost estimates were adjusted to 2024 U.S. dollars using the Consumer Price Index for Medical Care, where applicable.

### Cost-of-Illness Estimation

We estimated per-patient influenza costs by age category, including outpatient medical visits, inpatient hospitalizations, prescription medications, over-the-counter medications, and productivity losses. Productivity losses were estimated using age-specific median weekly earnings from the BLS, adjusted by labor force participation rates for each age group. Lost earnings were applied to symptomatic influenza cases based on published estimates of work absence associated with influenza illness.

Total seasonal costs for the 2023–24 influenza season were calculated by multiplying per-patient costs by CDC estimates of the number of symptomatic illnesses, medical visits, hospitalizations, and deaths by age category.

### Statistical Analysis

To examine the relationship between vaccination coverage and influenza severity, we estimated fixed-effects panel regressions relating the natural logarithm of hospitalization rates and mortality rates to the natural logarithm of influenza vaccination rates. The analysis covers influenza seasons from 2010–11 through 2023–24 and is conducted at the age-group–season level.

Separate models were estimated for (1) all adults aged ≥18 years and (2) adults aged ≥50 years, reflecting differences in influenza risk profiles and vaccine effectiveness across age groups. Age-group fixed effects were included to control for time-invariant differences across cohorts, while seasonal variation in influenza severity is captured through the panel structure. Statistical significance was assessed at the 5% level.

Medical visits and symptomatic illnesses were not directly estimated via regression. Instead, these outcomes were derived from hospitalization estimates using scalars consistent with CDC burden modeling methodologies, which infer non-hospitalized outcomes from laboratory-confirmed hospitalization data.

### Scenario Analyses

Scenario analyses were conducted to assess how alternative vaccination rates would have affected influenza outcomes and economic costs during the 2023–24 season. Two counterfactual scenarios were evaluated: (1) vaccination rates equal to the highest age-specific coverage observed between the 2010–11 and 2023–24 seasons, and (2) vaccination rates equal to the lowest observed coverage during the same period.

Regression coefficients were applied to estimate changes in mortality and hospitalization rates under each scenario. Corresponding changes in medical visits, symptomatic illnesses, healthcare costs, productivity losses, and deaths were then calculated using the cost-of-illness framework.

### The Estimated Annual Economic Cost of Influenza

Numerous studies have quantified the economic burden of influenza. These include costs associated with influenza-related outpatient visits, prescription and over-the-counter medication purchases, hospitalizations, and lost productivity from missed work. The cost estimates vary depending on the demographics examined and the comprehensiveness of the costs considered.

Hu et al. (2024) performed a retrospective cohort study of adults aged 18–49, 50–64, and _≥_65 years.^5^ The analysis found that mean per-patient hospitalization costs for influenza, including associated emergency department visits, ranged from $11,384 to $12,896 during the 2022–23 flu season. These costs varied by age group and season severity. For example, during the high-severity 2017–18 season, mean per-patient hospitalization costs ranged from $12,556 to $14,494. Medical visit–related costs ranged from $612 to $865 in 2017–18 and from $1,147 to $1,650 in 2022–23.

Langer et al. (2024) conducted a systematic literature review on the annual burden of influenza, including its economic impact, across multiple countries.^6^ In the United States, per-patient hospitalization costs ranged from $18,770 to $31,237 across the studies reviewed.

A literature review by de Courville et al. (2022) examined the economic burden of influenza among adults aged 18–64 years.^7^ Reported hospitalization costs ranged from $7,067 for adult patients with uncomplicated influenza to $38,662 for adults aged 45–59 years, with higher per-patient costs in the 45–59 age group compared with those aged 18–45. The studies reviewed also indicated that indirect costs (e.g., lost productivity) accounted for 83% to 99% of total costs for the 18 to 64 year old age group.

Federici et al. (2018) conducted a systematic literature review of healthcare costs associated with influenza in high-income countries.^8^ For the United States, studies estimated inpatient hospitalization costs ranging from $4,526 to $38,662 per patient, depending on age and clinical severity. Reported drug costs ranged from $5 for over-the-counter medications to $60 for prescription medications.

Putri et al. (2018) estimated the average annual economic burden of seasonal influenza based on the 2015 U.S. population.^9^ They reported a total annual burden of $11.2 billion, comprising $3.2 billion in direct medical costs and $8.0 billion in indirect costs, including an estimated 20.1 million lost “days of productivity.”

Aggarwal et al. (2018) evaluated hospitalization costs and length of stay for seasonal influenza using the 2015 National Inpatient Sample.^10^ The mean age of hospitalized patients was 61.2 years, the average length of stay was 4.7 days, and the mean hospitalization charges were $35,248.

Noelle-Angélique et al. (2007) estimated the medical and indirect costs associated with influenza based on the 2003 U.S. population.^11^ They found that annual influenza epidemics accounted for 3.1 million hospitalization days, $10.4 billion in direct medical costs, and $16.3 billion in lost earnings. When the statistical value of life was included, the total economic burden was estimated at $87.1 billion, including approximately 44 million lost days of productivity. In a 2022 systematic review of the productivity losses due to flu, Zumofen et al. found that the average number of lost workdays ranged from 0.5 to 5.3.^12^ Notably, the authors also found “studies evaluating vaccination status generally demonstrate that smaller proportions of vaccinated employees missed time from work due to influenza/ILI.”

We synthesize findings from Hu et al. (2024), Federici et al. (2018), and Noelle-Angélique et al. (2007), augmented with contemporary income and cost data, to estimate annual per-patient influenza costs that include medical visits, hospitalizations, prescription drugs, over-the-counter drugs, and productivity losses. Hu et al. (2024) is used as the primary source because it provides the most recent estimates of influenza-related healthcare costs. Since Hu et al. (2024) does not provide estimates of lost work hours and drug costs, we rely on Noelle-Angélique et al. (2007) for estimates of lost work hours and on Federici et al. (2018) for estimates of prescription and over-the-counter drug costs. These cost estimates are updated to 2024 prices using the Consumer Price Index for Medical Care and are presented in Table 1.

**Table 1.** Annual Per-Patient Influenza Costs Medical Visits, Prescription Drugs, Over-the-Counter Drugs, Hospitalization, and Lost Earnings, By Age Category.

| Per Patient Costs |  |  |  |
| --- | --- | --- | --- |
|  | 18-49 | 50-64 | 65+ |
| Medical Visit Costs | \$964 | \$705 | \$798 |
| Prescription Drug Costs | \$83 | \$83 | \$83 |
| Over-the-Counter Drug Costs | \$11 | \$11 | \$11 |
| Inpatient Hospital Costs | \$12,993 | \$14,081 | \$12,498 |
| Median Weekly Earnings | \$1,112 | \$1,246 | \$1,154 |
| Labor Participation Rate | 83.6% | 74.1% | 17.9% |
| Participation Adj. Median Weekly Earnings | \$930 | \$923 | \$206 |

The medical visit costs for each age category are calculated as the average of the reported costs for a standalone emergency department visit in the 2022–23 flu season from Hu et al. (2024) and the average uninsured primary care visit cost reported by AdventHealth.^13^ Per-patient prescription and over-the-counter drug costs are based on the average of values reported by Federici et al. (2018). Inpatient hospital costs for each age category are taken from Hu et al. (2024) for the 2022–23 flu season. Median weekly earnings are obtained from the Bureau of Labor Statistics (BLS) for 2024Q2.^14^ They are adjusted by the labor force participation rate for each age category, as reported by the BLS.

### Documenting The Annual Cost of Influenza

The total economic burden of the 2023–24 influenza season can be estimated by combining these per-patient cost estimates with CDC annual estimates of the number of symptomatic influenza illnesses, medical visits, hospitalizations, and influenza-associated deaths. The CDC reports these outcomes for the overall population and by age category. The 2023–24 flu season estimates used in this analysis are presented in Table 2.

**Table 2.** Patients Experiencing Flu Illness, Medical Visits, Hospitalization, and Death, By Age Category 2023-24 Flu Season.

| 2023-24 Flu Season |  |  |  |  |
| --- | --- | --- | --- | --- |
| Patients Experiencing Flu Illness, Medical Visits, Hospitalization, and Death |  |  |  |  |
|  | 18-49 | 50-64 | 65+ | 18+ |
| Population | 140,303,491 | 62,531,182 | 59,248,361 | 262,083,034 |
| Patients w/Symptomatic Illness | 16,351,383 | 9,042,884 | 2,610,181 | 28,004,448 |
| Patients w/Medical Visits | 6,050,012 | 3,888,440 | 1,461,701 | 11,400,153 |
| Patients Hospitalized | 91,780 | 95,897 | 237,289 | 424,966 |
| Patient Mortality | 2,705 | 5,701 | 19,038 | 27,444 |

Applying the per-patient cost estimates to the influenza incidence data yields an estimate of the economic costs associated with the 2023–24 influenza season. For adults aged _≥_18 years, the 2023–24 season was associated with 27,444 deaths, approximately $16 billion in direct healthcare costs, and more than $13 billion in lost productivity, for a total economic burden of nearly $29 billion (Table 3). These estimates are broadly consistent with previous cost-of-illness analyses.

**Table 3.** Estimated Total Influenza Costs 2023-24 Flu Season By Age Category Vaccination Remains the Primary Prevention Strategy.

| Total Costs: 2023-24 Flu Season |  |  |  |  |
| --- | --- | --- | --- | --- |
|  | 18-49 | 50-64 | 65+ | 18+ |
| Medical Visit Costs | \$5,831,597,163 | \$2,742,581,368 | \$1,166,970,849 | \$9,741,149,380 |
| Prescription Drug Costs | \$234,105,901 | \$150,463,627 | \$56,560,686 | \$441,130,214 |
| Over-the-Counter Drug Costs | \$155,206,462 | \$82,949,661 | \$22,138,767 | \$260,294,890 |
| Hospital Costs | \$1,192,482,577 | \$1,350,290,031 | \$2,965,585,416 | \$5,508,358,024 |
| *Healthcare Costs* | *\$7,413,392,102* | *\$4,326,284,686* | *\$4,211,255,719* | *\$15,950,932,507* |
| *Productivity Losses* | *\$8,708,585,317* | *\$4,239,555,703* | *\$65,793,973* | *\$13,013,934,993* |
| **Total Flu Healthcare Costs and Productivity Losses** | | | | **\$28,964,867,500** |

Annual influenza vaccination is the primary prevention strategy, and substantial evidence supports its impact on reducing adverse outcomes.^15^ Interim 2024–25 vaccine-effectiveness data indicate reductions of 36%–54% in outpatient illness and 41%–55% in hospitalization among vaccinated adults.^16^ Vaccination has also been associated with reduced risk of intensive care unit admission and death among hospitalized adults.^17^ Meta-analyses and CDC modeling further demonstrate that vaccination not only lowers the risk of infection but also mitigates illness severity, healthcare utilization, and productivity losses.^18^ In modeling analyses, influenza vaccination prevented an estimated 1.9 million illnesses in the 2021–22 season, illustrating how higher uptake correlates with improved outcomes across populations.^19^ Despite public health efforts, influenza vaccination coverage remains below national targets. Table 4 and Figure 1 summarize vaccination rates by age cohort from the 2010–11 through the 2023–24 flu seasons.

**Figure 1.**
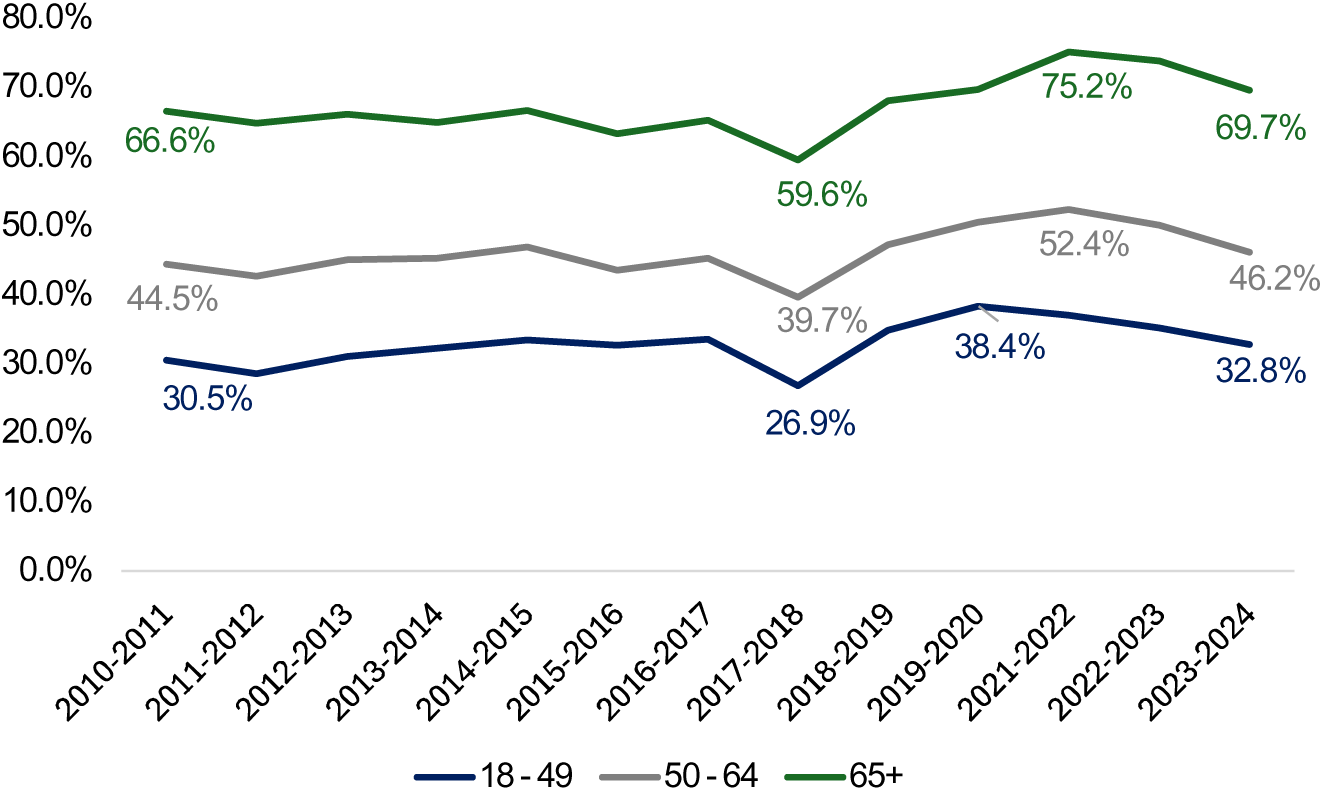
Influenza Vaccination Rates By Age Category, 2010-11 Flu Season Through 2023-24 Flu Season.

**Table 4.** Influenza Vaccination Rates By Age Category, 2010-11 Flu Season Through 2023-24 Flu Season.

| Vaccination Rates |  |  |  |
| --- | --- | --- | --- |
| Flu Season | 18 - 49 | 50 - 64 | 65+ |
| 2010-2011 | 30.5% | 44.5% | 66.6% |
| 2011-2012 | 28.6% | 42.7% | 64.9% |
| 2012-2013 | 31.1% | 45.1% | 66.2% |
| 2013-2014 | 32.3% | 45.3% | 65.0% |
| 2014-2015 | 33.5% | 47.0% | 66.7% |
| 2015-2016 | 32.7% | 43.6% | 63.4% |
| 2016-2017 | 33.6% | 45.4% | 65.3% |
| 2017-2018 | 26.9% | 39.7% | 59.6% |
| 2018-2019 | 34.9% | 47.3% | 68.1% |
| 2019-2020 | 38.4% | 50.6% | 69.8% |
| 2021-2022 | 37.1% | 52.4% | 75.2% |
| 2022-2023 | 35.2% | 50.1% | 73.9% |
| 2023-2024 | 32.8% | 46.2% | 69.7% |

Table 4 and Figure 1 illustrate variation in influenza vaccination rates across age cohorts and over time. Vaccination coverage increases as the 65+ age category has a higher vaccination rate than the 50–64 age category, which in turn has a higher vaccination rate than the 18–49 age category. Following a steady recovery after the 2017–18 low point, vaccination rates have again declined across all cohorts. Rates in 2023–24, although higher than in 2017–18, remain substantially below prior peak levels. This downward trend reinforces the need for renewed emphasis on economic evaluation and evidence-based interventions to sustain and improve vaccine uptake.

These trends underscore the importance of a comprehensive cost-of-illness analysis that quantifies both the clinical and economic burden of influenza in the United States. Understanding how vaccination rates influence healthcare utilization and associated costs is essential for assessing the value of preventive interventions and for guiding evidence-based policy.

### The Empirical Relationship Between Vaccination Rates and Flu Illness Severity

To assess how alternative vaccination rates could affect influenza outcomes and costs, we conducted two separate panel-data regressions with fixed effects. These regression models relate the natural logarithms of mortality and hospitalization rates to the natural logarithm of vaccination rates, using historical CDC influenza data. Our a priori hypothesis was that higher vaccination rates would be associated with lower mortality and hospitalization rates.

Rather than estimating the number of medical visits and symptomatic illnesses directly via panel regression, we derive these values from the estimated hospitalization rates. This approach parallels the CDC’s methodology, which uses “laboratory-confirmed flu-associated hospitalization rates obtained from the Influenza Hospitalization Surveillance Network” to estimate hospitalization rates and then infers medical visits and symptomatic illnesses from established relationships between hospitalizations and these outcomes.

The data cover influenza seasons from 2010–11 through 2023–24 and are analyzed by age group. Because influenza severity and risk profiles differ between younger and older adults, we estimate models both for all three adult age cohorts (18–49, 50–64, and ≥65 years) and for older adults only (50–64 and ≥65 years). Table 5 presents the underlying CDC data on symptomatic illnesses, medical visits, hospitalizations, and deaths.

**Table 5.**
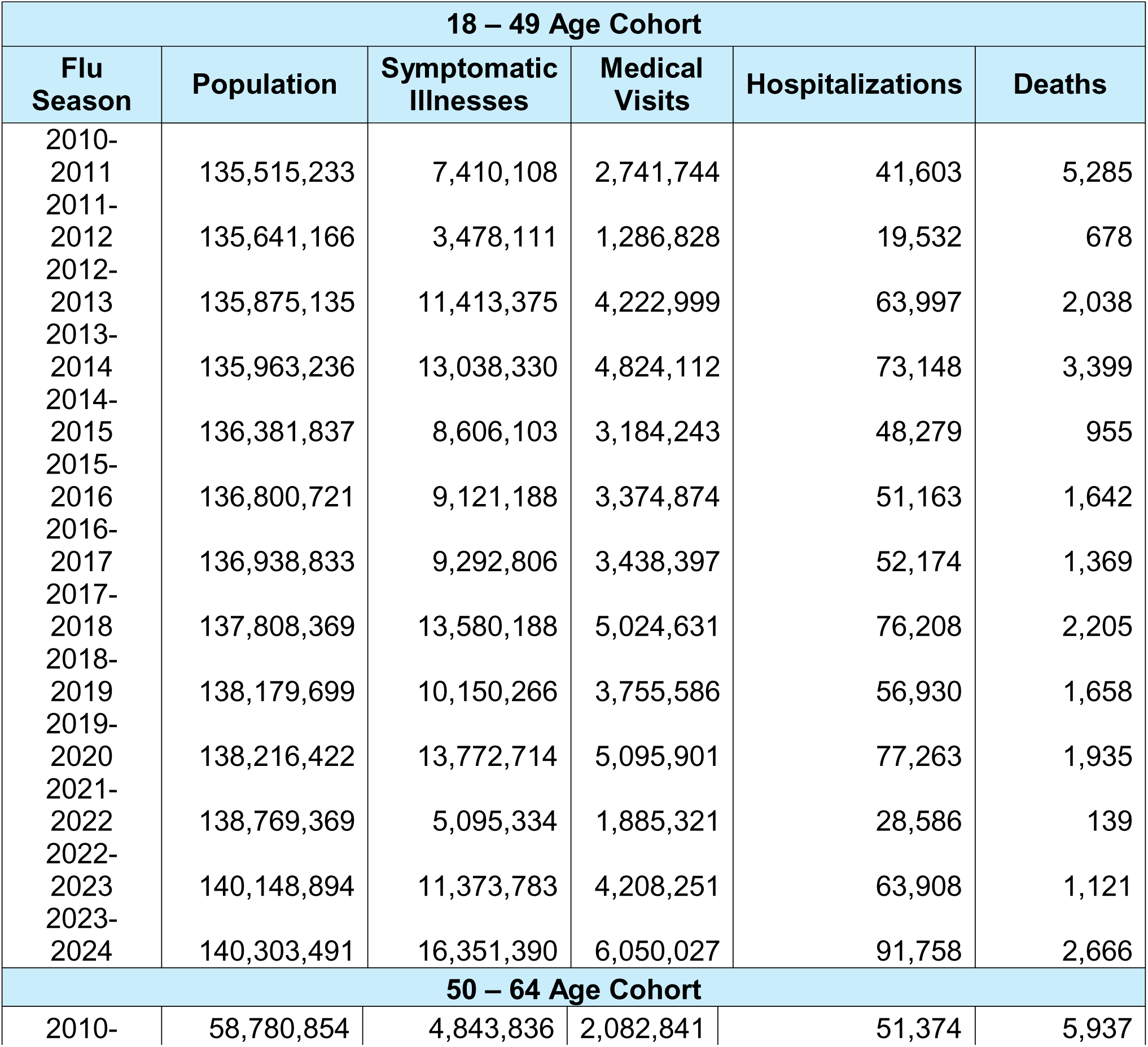
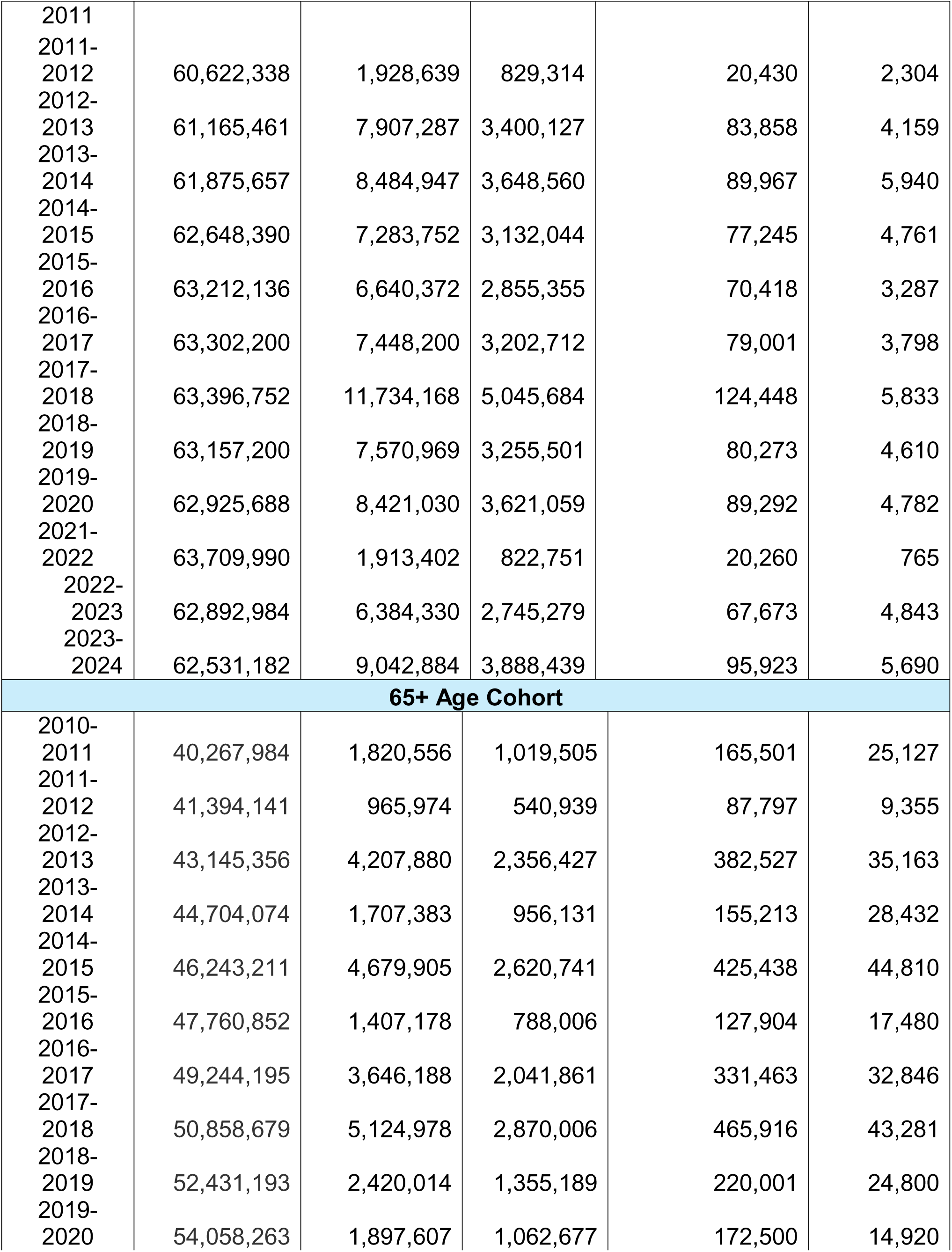
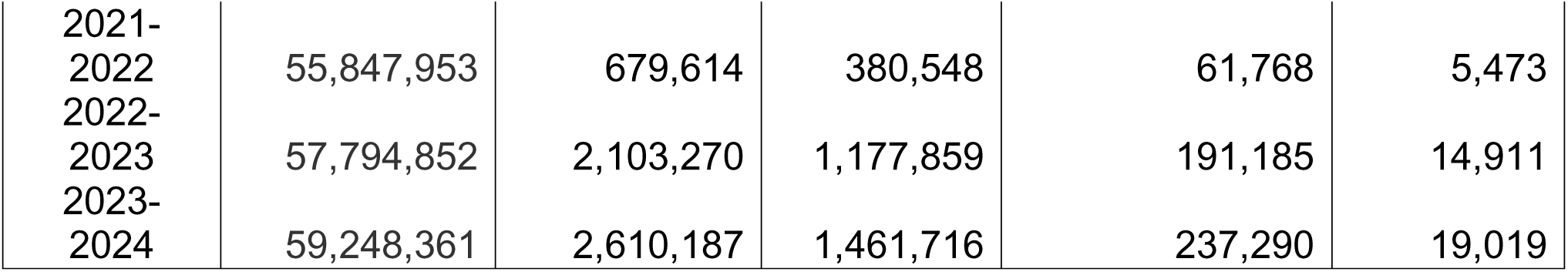
Influenza Mortalities, Hospitalizations, Medical Visits, and Symptomatic Illnesses By Age Category, 2010-11 Flu Season Through 2023-24 Flu Season.

**Table 6.** Panel Data Regression Results 50+ and 18+ Age Categories.

| Panel Regression Results: 50+ Age Categories |  |  |  |  |
| --- | --- | --- | --- | --- |
|  | Hospitalizations |  | Mortality |  |
|  | *Coefficients* | *Standard Error* | *Coefficients* | *Standard Error* |
| Intercept | (9.5099) | 1.2798 | (13.3973) | 1.2425 |
| Vaccination Rate | (3.4160) | 1.6376 | (4.8040) | 1.5900 |
| 65+ | 2.6411 | 0.6548 | 3.7245 | 0.6357 |
| Adjusted R Square | 0.6251 |  | 0.7836 |  |
| F-Statistic | 21.8447 |  | 46.2737 |  |

Data are as a percentage of the relevant population
| Panel Regression Results: 18+ Age Categories |  |  |  |  |
| --- | --- | --- | --- | --- |
|  | Hospitalizations |  | Mortality |  |
|  | *Coefficients* | *Standard Error* | *Coefficients* | *Standard Error* |
| Intercept | (9.4396) | 1.2529 | (16.0084) | 1.5395 |
| Vaccination Rate | (1.4186) | 1.1145 | (4.1000) | 1.3694 |
| 65+ | 3.3665 | 0.8271 | 6.6159 | 1.0162 |
| 50 - 64 | 1.4795 | 0.4332 | 3.1573 | 0.5323 |
| Adjusted R Square | 0.7678 |  | 0.8470 |  |
| F-Statistic | 42.8819 |  | 71.1110 |  |

The regression analysis demonstrates that influenza vaccination rates are significantly and inversely associated with hospitalization and mortality among adults aged 50 and older (p < 0.05). When all adult cohorts (18+) are included, vaccination remains a statistically significant predictor of lower mortality rates but not hospitalization rates. These findings confirm that vaccination mitigates mortality risk across all adults and reduces severe illness among individuals aged 50 and older.

Given the strong inverse relationship between vaccination and disease severity, even modest improvements in coverage could yield substantial cost savings through reduced total healthcare expenditures and productivity losses. The reverse is also true. Scenario analyses can illustrate the impact of vaccines by quantifying how alternative vaccination rates would affect influenza outcomes, including mortality, hospitalizations, medical visits, and symptomatic illness.

To evaluate how changes in vaccination rates could affect these outcomes, we examine how alternative vaccination rates could have altered the costs associated with the 2023-24 flu season. Since higher vaccination rates are associated with lower mortality among adults aged ≥18 years, we use panel regression estimates for this age group. For hospitalizations, higher vaccination rates are only associated with lower hospitalization rates among adults aged ≥50 years; therefore, we use the estimates for adults aged ≥50 years. We then apply a scalar, derived from the 2023–24 flu season data, that relates hospitalizations to medical visits and symptomatic illnesses to project the corresponding changes in those outcomes.

The first scenario evaluates how the economic burden of the 2023–24 flu season would have changed if vaccination rates in each age category had matched their peak levels observed between the 2010–11 and 2023–24 seasons. These peak vaccination rates were 38.4% for adults aged 18–49, 52.4% for adults aged 50–64, and 75.2% for adults aged _≥_65 years. Table 7 presents the results. Under this scenario, the total economic burden of the 2023–24 season would have been approximately $2.9 billion lower, with more than 8,000 fewer influenza-related deaths.

**Table 7.**
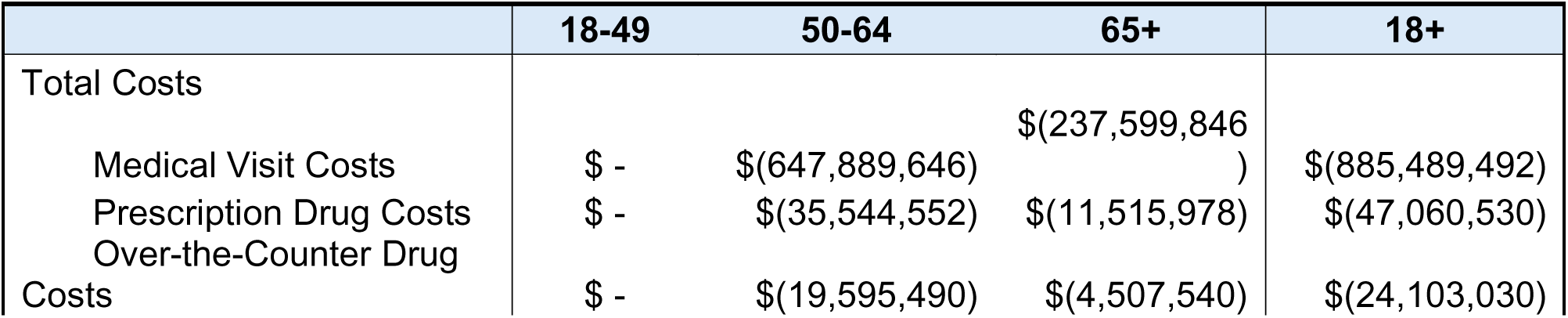
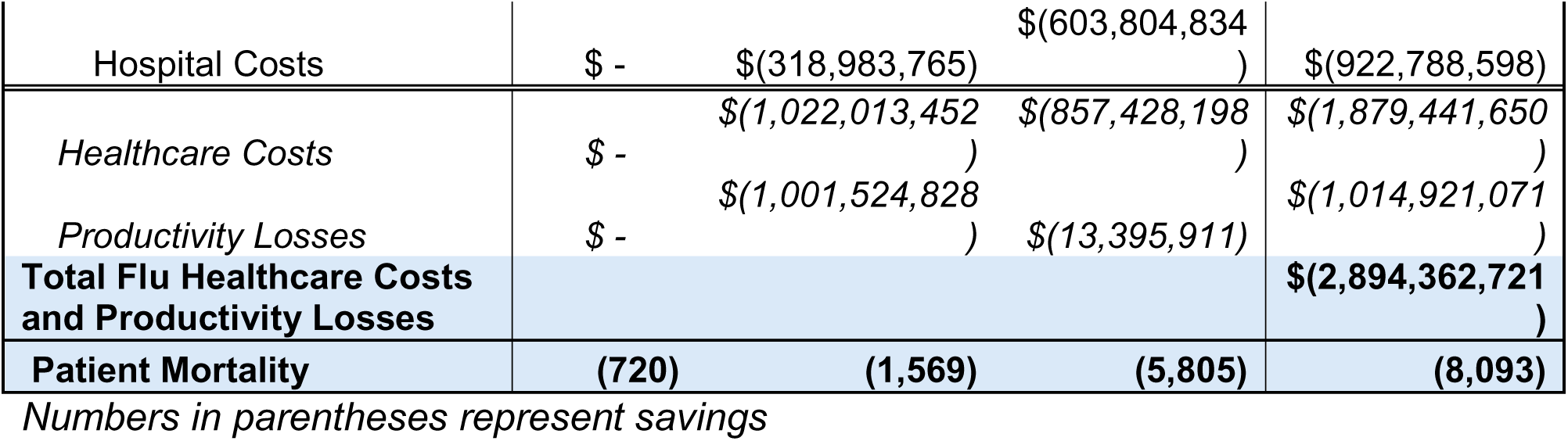
Changes in Economic Costs and Mortalities Associated with 2023-24 Flu Season Had Vaccination Rates Equaled Highest Vaccination Rates Since 2010-11 Flu Season.

The second scenario evaluates how the economic burden of the 2023–24 flu season would have changed if vaccination rates in each age category had instead matched their lowest levels observed between the 2010–11 and 2023–24 seasons. These lowest vaccination rates were 26.9% for adults aged 18–49, 39.7% for adults aged 50–64, and 59.6% for adults aged _≥_65 years. As shown in Table 8, under this scenario the total economic burden of the 2023–24 season would have been approximately $6.6 billion higher, with nearly 25,000 additional influenza-related deaths.

**Table 8.**
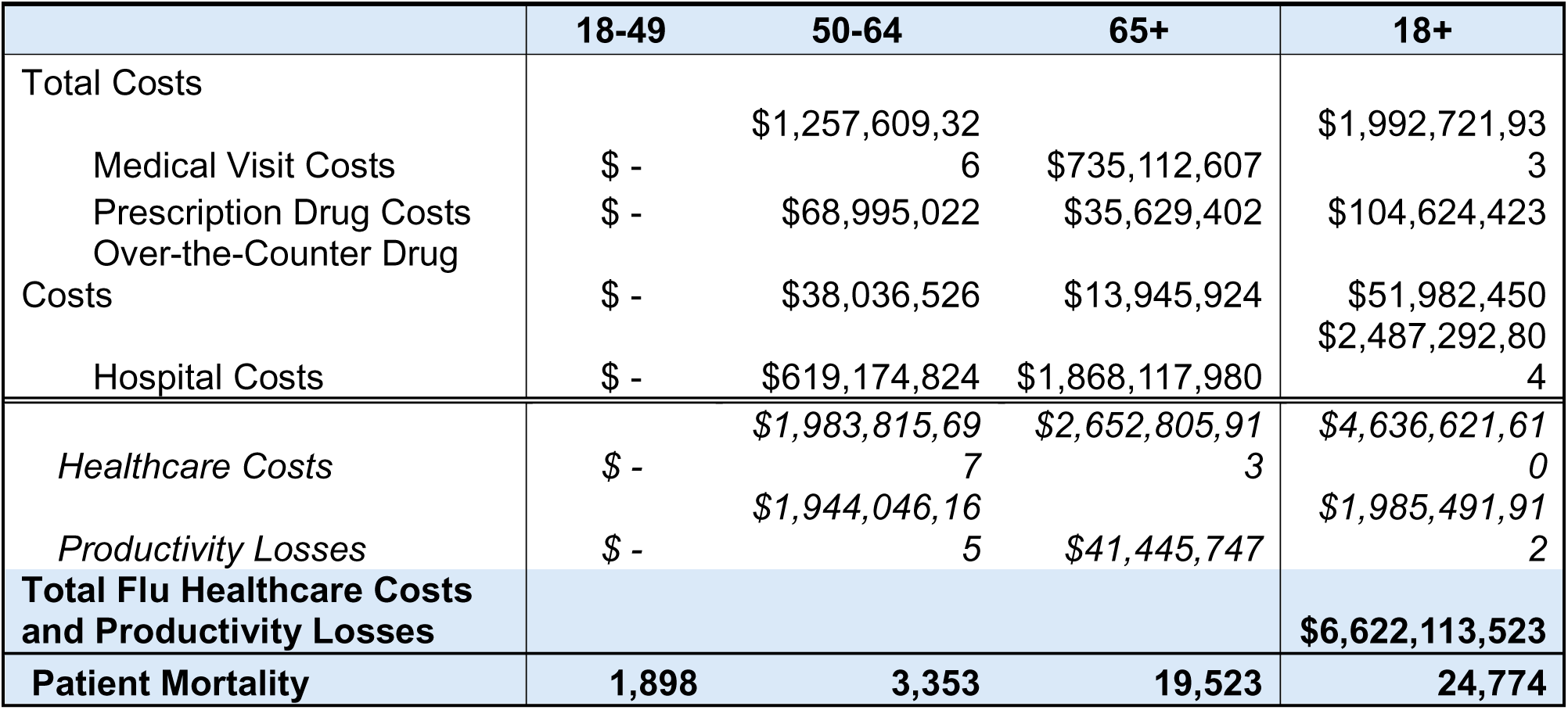
Changes in Economic Costs and Mortalities Associated with 2023-24 Flu Season Had Vaccination Rates Equaled Lowest Vaccination Rates Since 2010-11 Flu Season.

These scenarios illustrate that changes in vaccination rates can meaningfully affect both the economic costs and mortality associated with influenza. Hence, improving coverage reduces economic and humanistic burdens, whereas declining coverage substantially increases them.

## Discussion

This analysis confirms that seasonal influenza continues to impose a substantial clinical and economic burden on adults in the United States. For the 2023–24 influenza season, we estimate that influenza was associated with nearly $29 billion in combined direct healthcare costs and productivity losses among adults, alongside 27,444 deaths. Direct medical costs, including outpatient visits, hospitalizations, prescription drugs, and over-the-counter medications, accounted for approximately $16 billion, while indirect costs from lost productivity exceeded $13 billion. These findings are broadly consistent with earlier cost-of-illness studies, while updating estimates to reflect current utilization patterns and price levels.

Our results highlight differences across adult age groups. Adults aged 65 years and older account for a disproportionate share of hospitalizations and deaths, reflecting their higher clinical vulnerability. At the same time, adults aged 18 to 64 contribute heavily to the productivity component of the burden of illness, given their higher labor force participation and median earnings. This dual pattern underscores that influenza has meaningful implications for both healthcare utilization and economic performance among adults.

The panel-data regression analysis demonstrates a statistically significant inverse association between influenza vaccination rates and both hospitalization and mortality among adults aged 50 years and older. When all adults aged ≥18 years are included, higher vaccination coverage remains significantly associated with lower mortality, although the association with hospitalization rates is no longer statistically significant. These findings are consistent with the well-established evidence that influenza vaccination most strongly reduces the risk of severe outcomes, particularly death, among older and higher-risk adults, while still conferring protection across the broader population.

The scenario analyses further quantify the potential implications of changes in adult vaccination coverage. If age-specific vaccination rates in 2023–24 had matched their historical peak levels observed between 2010–11 and 2023–24, the total economic burden of influenza would have been reduced by approximately $2.9 billion, and more than 8,000 deaths would have been averted in a single season. In contrast, if coverage had fallen to the lowest levels observed during this period, the total burden would have increased by about $6.6 billion, resulting in nearly 25,000 additional deaths. These scenarios, while based on modeled associations, illustrate the magnitude and direction of changes that could plausibly arise from relatively modest shifts in vaccination uptake. Taken together, these results reinforce the central role of vaccination in influenza control.

Several limitations should be noted. First, the analysis relies on CDC burden estimates, which are model-based and subject to uncertainty in surveillance and attribution. Second, the cost inputs are drawn from multiple studies conducted in different years and populations and then updated to 2024 prices. However, this approach is standard in cost-of-illness work; it introduces potential heterogeneity that cannot be fully resolved.

## Conclusion

This study provides an updated cost-of-illness assessment of seasonal influenza among U.S. adults, integrating recent CDC burden estimates with contemporary cost data and panel regression analyses of vaccination and outcomes. Higher vaccination coverage, particularly among adults aged _≥_50 years, is associated with substantially lower hospitalization and mortality rates, and scenario analyses suggest that realistic improvements or declines in coverage could shift the annual economic burden by several billion dollars and alter influenza-related deaths by tens of thousands.

These findings reinforce the value of adult influenza vaccination as both a public health and an economic intervention. Policies improving adult influenza vaccination uptake should therefore be viewed as a central component of strategies to enhance population health and protect the resilience of our healthcare system.

## Data Availability

All data produced in the present study are available upon reasonable request to the authors

